# Fecal biomarkers of environmental enteric dysfunction and the gut microbiota of rural Malawian children: an observational study

**DOI:** 10.1101/2021.01.19.21250144

**Authors:** David Chaima, Harry Pickering, John D. Hart, Sarah E. Burr, Kenneth M. Maleta, Khumbo Kalua, Robin L. Bailey, Martin J. Holland

## Abstract

Environmental enteric dysfunction (EED) is a subclinical condition of the gut characterized by changes in morphology and function with underlying chronic inflammatory responses. This study characterized composition and diversity of the gut microbiota in rural Malawian children with and without signs of EED. Fecal samples were collected from children aged 1-59 months. Neopterin, myeloperoxidase and alpha-1 antitrypsin concentrations were quantified by ELISA and combined to form a composite EED score using principal component analysis. DNA was extracted from fecal samples and V4-16S rRNA sequencing was used to characterize the gut microbiota. The concentrations of all three biomarkers decreased with increasing age, which is consistent with other studies of children living in similar low-income settings. *Firmicutes, Bacteroidetes, Proteobacteria* and *Actinobacteria* were the dominant phyla while *Faecalibacterium* and *Bifidobacterium* were the most prevalent genera. Increased alpha diversity was associated with a reduction in neopterin concentration. Microbiota composition was associated with the composite EED score. Increased abundance of *Succinivibrio* was associated with reduced composite EED scores.

**Highlights:**

- In Malawian children, fecal concentrations of myeloperoxidase, alpha-1 antitrypsin and neopterin decreased with age
- A marginal negative association was found between alpha diversity of the gut microbiota and fecal neopterin concentration
- A higher abundance of *Succinivibrio* was found in children with lower concentrations of biomarker of environmental enteric dysfunction
- Fatty acid biosynthesis, tetrapyrrole biosynthesis and pyrimidine nucleotide degradation pathways were associated with environmental enteric dysfunction biomarker score

## Introduction

The gut microbiota is essential in maintaining normal physiological and metabolic processes. Changes in the gut microbiota composition and function have been associated with a number of conditions including environmental enteric dysfunction (EED)^1,2^. EED is characterized by partial villous atrophy, impaired enterocyte tight junctions and underlying chronic inflammatory responses. The specific cause of EED has not yet been clearly established, however, one model suggests that ingestion of contaminated food or water changes the composition and function of the gut microbiota, resulting in intestinal inflammation and morphological changes to the intestinal epithelium thereby affecting normal gut function^2,3^. EED affects more than two-thirds of children aged between 0-5 years living in low-income settings^4,5^ and has been linked with growth faltering and reduced efficacy of oral vaccines^6^.

Confirmation of a diagnosis of EED requires visualization of the intestinal epithelium using endoscopy or biopsy, however, these are invasive tests that would be unethical to perform in asymptomatic children. The urinary lactulose: mannitol (L:M) ratio test has been extensively used in the past as a non-invasive test to diagnose EED however, the test is inconsistent; procedural details, including fasting prior to ingestion, sugar dosage, time of urine collection and method of detection, vary between studies making comparison of results difficult. Leakage of urine sample from pediatric urine bags and urine contamination by stool are other additional challenges in infants. Potential biomarkers that can easily be quantified in fecal samples and blood using ELISA techniques have been tested as biomarkers of EED^7-11^. These biomarkers are thought to measure the various processes involved in EED, such as intestinal epithelial damage, intestinal inflammation, intestinal permeability, microbial translocation and systemic inflammation^2^. Myeloperoxidase (MPO), alpha-1 antitrypsin (AAT) and neopterin (NEO) are three potential biomarkers that have been evaluated for use in EED diagnosis, although available data suggests that no single biomarker is sufficient to indicate EED^6,12^. Accordingly, a multi-site, longitudinal study by Kosek *et al*.^13^ combined the absolute measured concentrations of AAT, MPO and NEO into a single metric, using principal component analysis, to form a composite EED score. They then explored the relationship between the composite EED score and linear growth in infants from resource limited settings. That study reported an association between higher composite EED scores and growth faltering. In the present study, we used fecal samples collected from rural Malawian children to characterize the fecal microbiota defined by V4 16S rRNA gene sequencing and explored associations with EED indicated by fecal levels of MPO, AAT and NEO.

## Results

### Distribution of individual biomarker concentration and the composite EED score

One hundred and two baseline samples were analyzed by 16S rRNA sequencing as detailed elsewhere^14^. Of these, 75 had sufficient sample remaining to quantify MPO, 69 for AAT and 64 for NEO. A total of 60 fecal samples had ELISA results for all 3 biomarkers. The age and sex of all 102 children who provided fecal samples for sequencing were not markedly different from those of the children who had sufficient sample to quantify all the 3 biomarkers or any of the 3 biomarkers (**Table 1**).

**Table 1:** Age and sex distribution of all children involved in 16S sequencing and those who had sufficient sample available for MPO, AAT and NEO assays

| Variable | All children (n=102) | All three biomarkers (n=60) | MPO (n=75) | AAT (n=69) | NEO (n=64) |
| --- | --- | --- | --- | --- | --- |
| Mean (SD) age in months | 23.0 (13.5) | 24.3 (13.8) | 23.3 (13.8) | 23.6 (14.1) | 24.0 (13.7) |
| Male sex, n (%) | 52 (51) | 34 (57) | 41 (55) | 38 (55) | 35 (55) |

The distribution of fecal biomarker concentrations was calculated in all available samples and compared by age. The concentration of all three biomarkers was skewed (**Figure 2**). A large proportion of children (81%) had elevated NEO levels relative to reference values (**Figure 2A)**. Ten percent of the children had elevated levels of fecal AAT (**Figure 2B**) and 38.9% (29/75) of children had elevated MPO levels (**Figure 2C**). The 3 biomarkers were combined to form a composite EED score as described and the range was plotted (**Figure 2D**). EED scores ranged from 0 (lowest score) to 3.46 (highest score) with a median and inter quartile range (IQR) of 2.26 (0.61).

**Figure 2.**
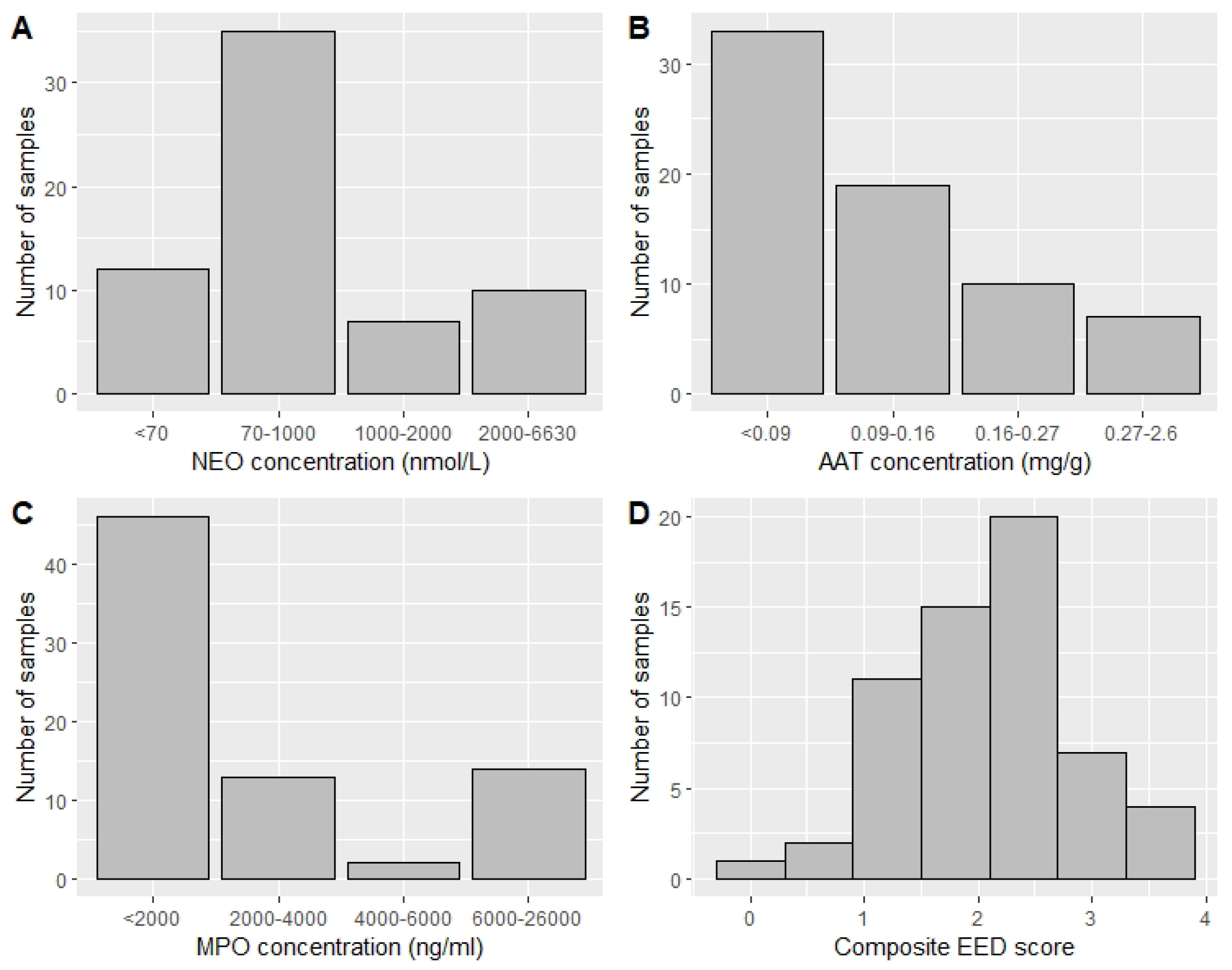
Distribution of individual biomarker concentrations and the composite EED score. (A) NEO, (B) AAT, (C) MPO and (D) composite EED score. Normal values for the individual biomarkers were based on values reported in the literature (NEO <=70 nmol/L, AAT <=0.27 mg/g and MPO <=2000 ng/ml)^6,12,13^.

### Effect of age on biomarker concentration

Age in months was categorized into 4 groups as follows: 0-12, 13-24, 25-36, and 37-60 and the Kruskal-Wallis test was used to test for the differences in biomarker concentration among the groups. Median fecal concentration for all the 3 biomarkers decreased with age (**Table 2**).

**Table 2:**
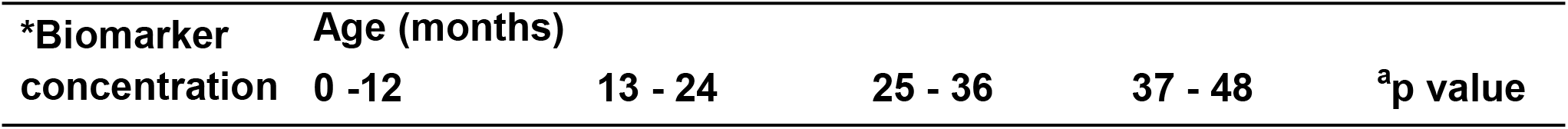
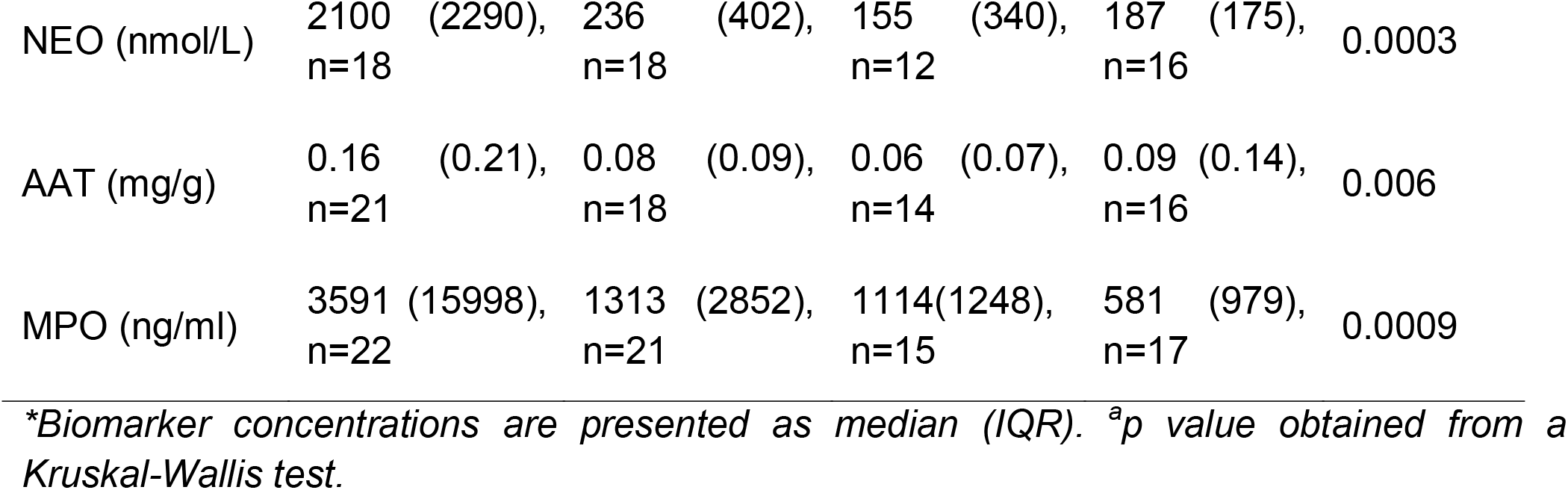
Median biomarker concentration by age.

### Gut microbiota and the effect of age

Characterization of the microbiota of this sample set has recently been described^14^. Briefly, the 102 samples generated a total of 1,685,792 reads with an average read depth per sample of 16,527±11,071. *Firmicutes, Bacteroidetes, Proteobacteria* and *Actinobacteria* were the most dominant phyla while *Faecalibacterium, Streptococcus, Bacteroides, Haemophilus, Subdoligranulum, Bifidobacterium* and *Escherichia-Shigella* were the most dominant genera. The mean (SD) Shannon index was 1.72 (0.45) and median (IQR) Simpson index was 0.75 (0.16).

In the present study, the effect of age on Shannon or Simpson diversity indices and the composition of the microbiota was explored among the 4 age groups. Shannon and Simpson indices were significantly lower in fecal samples from younger children compared to older children, indicating increasing diversity with age (**Figure 3A**, ANOVA p<0.001 and **Figure 3B**, Kruskal-Wallis p=0.004 respectively).

**Figure 3.**
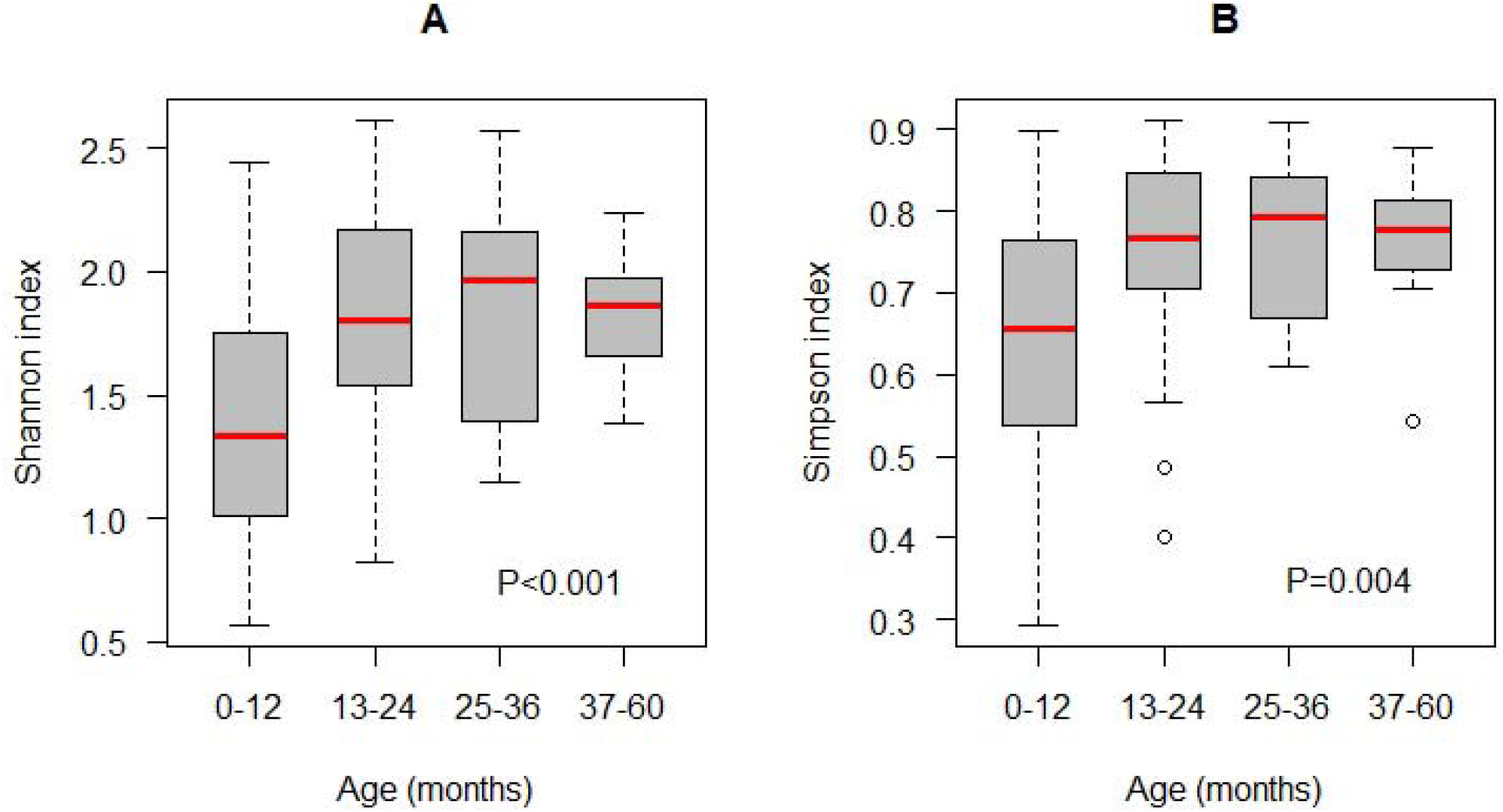
Comparison of the distribution of (A) Shannon and (B) Simpson indices by age group. p value for Shannon index (p<0.001) was obtained from ANOVA while p value for Simpson index (p=0.004) was obtained from the Kruskal-Wallis test.

Microbiota composition by age group was also examined using PERMANOVA. Comparisons of weighted and unweighted UniFrac distance showed significant differences in clustering of microbial communities by age (R^2^ =0.17, F=6.4, p=0.001 and R^2^=0.29, F=12.8, p=0.001 respectively) (**Figure 4**). Additionally, a comparison of relative abundance of microbial taxa by age showed that fecal samples from younger children (0-12 months) had a higher abundance of *Actinobacteria* and a lower abundance of *Bacteroidetes* than samples from older children (Kruskal-Wallis test p=0.01, **Figure 5A**). At the genus level, a higher abundance of *Bifidobacterium* and a lower abundance of genus *Faecalibacterium* were observed in younger children (0-12 months) compared to the older age groups (Kruskal-Wallis test p=0.01, **Figure 5B**).

**Figure 4.**
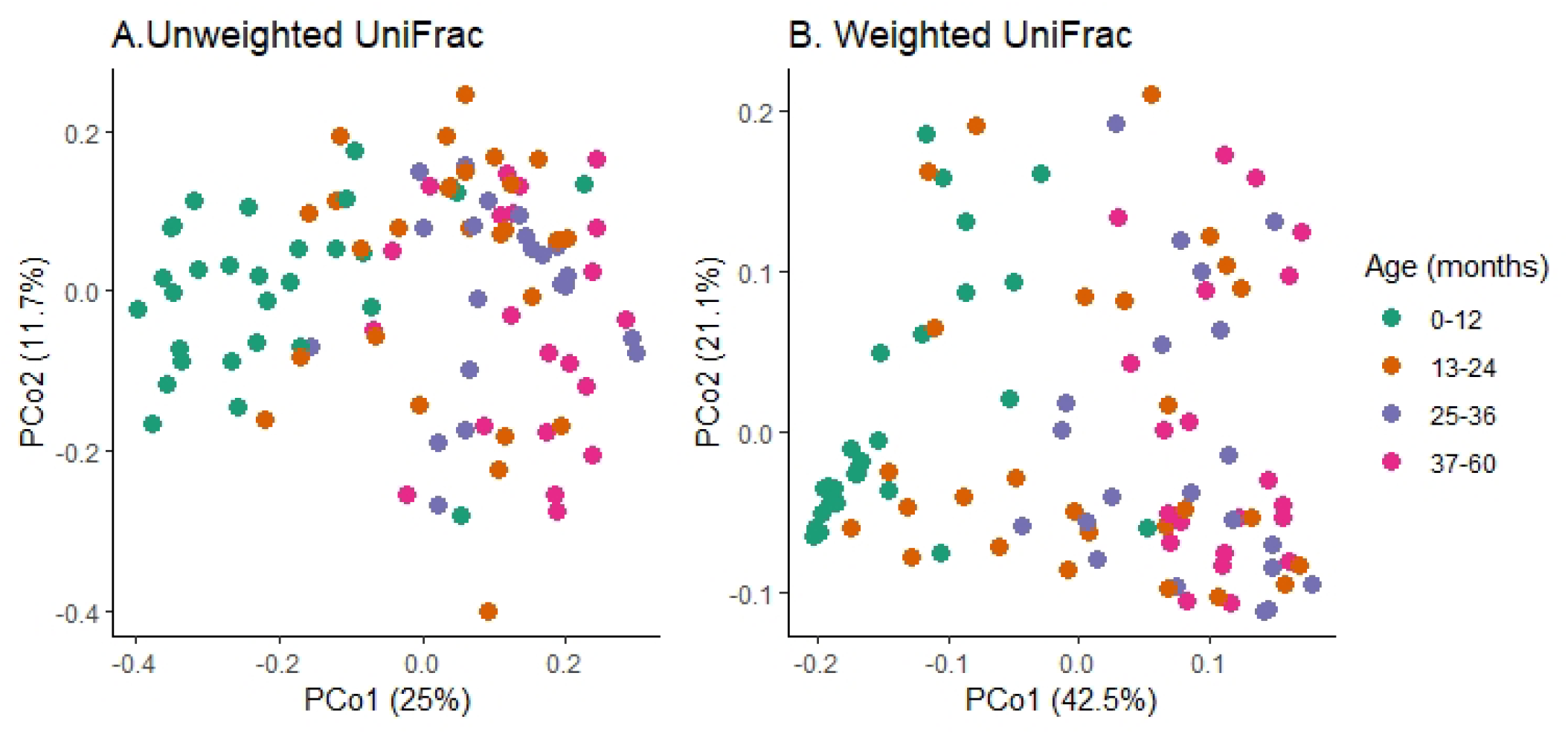
Overall microbiota composition differences by age. (A) PCoA plot of unweighted UniFrac distances (R^2^ =0.17, F=6.4, p=0.001) (B) PCoA plot of weighted UniFrac distances (R^2^=0.29, F=12.8, p=0.001).

**Figure 5.**
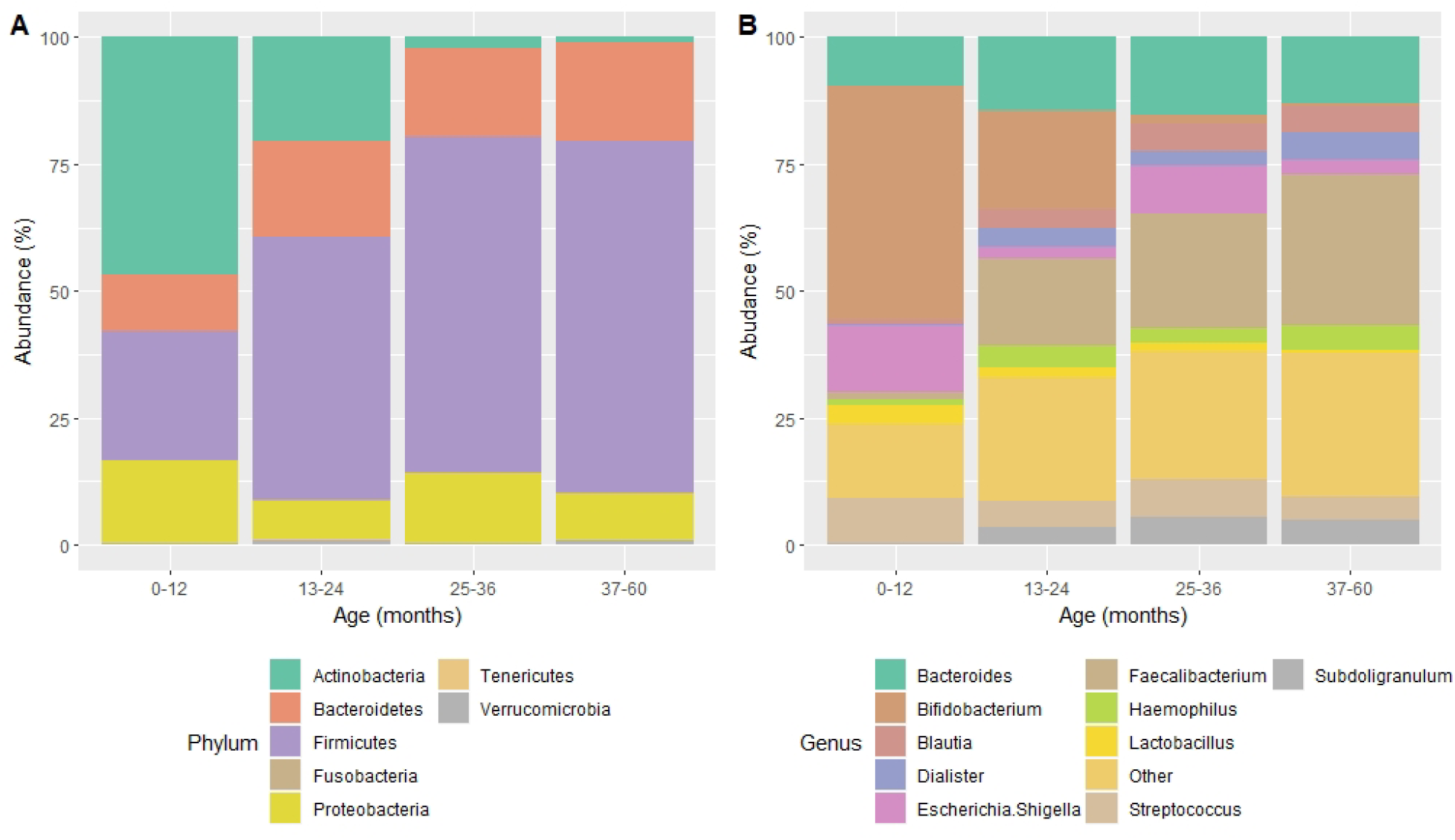
Relative abundance of taxa by age. (A) Relative abundance of phyla (B) Relative abundance of genera. The stacked bar plot shows the 10 most abundant genera; the remaining genera were grouped as “Other”.

### Associations between alpha diversity and fecal biomarkers of EED

Linear regression analysis adjusted for age and sex was used to examine the relationship between Shannon or Simpson diversity and fecal concentrations of NEO, AAT and MPO. There was no relationship between Shannon diversity and AAT or MPO concentrations, or between Simpson diversity and AAT or MPO concentrations. (**Table 3**). However, there were marginal negative associations between both Shannon and Simpson diversity, and NEO concentrations; increased Shannon or Simpson diversity was associated with a decrease in NEO concentration (**Table 3**). There was no association between Shannon or Simpson diversities and the composite EED scores (p=0.14 and P=0.33 respectively).

**Table 3:**
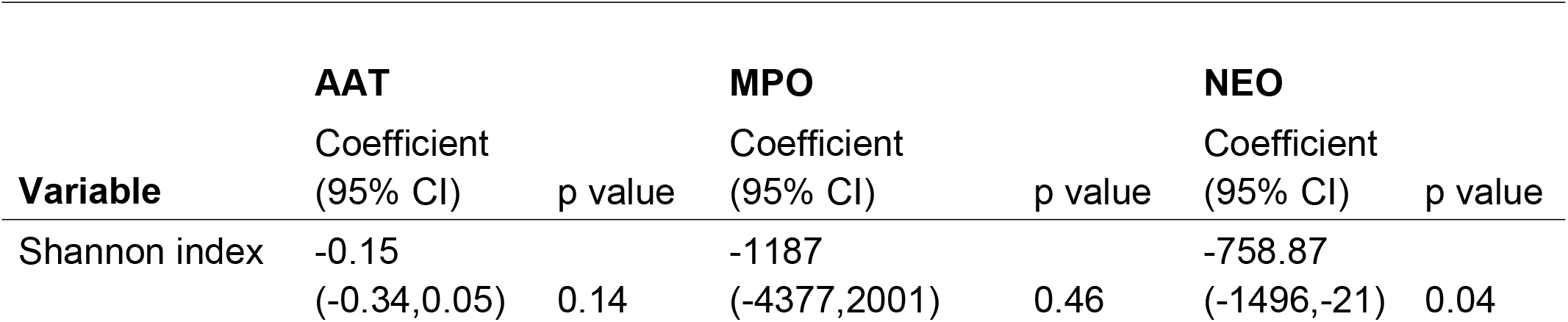
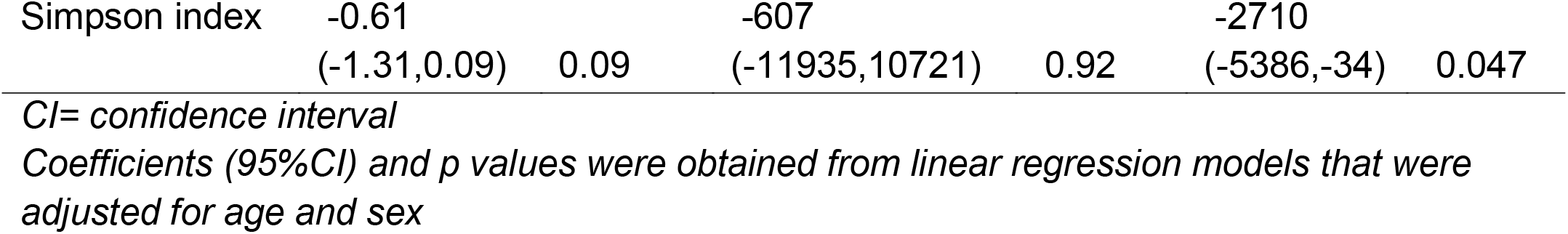
Relationship between alpha diversity indices and individual biomarkers of EED.

### Associations between fecal bacterial community structure and biomarkers of EED

PERMANOVA did not detect any differences in bacterial community composition between normal and elevated biomarker concentration for each of the 3 biomarkers (**Table 4**). However, PERMANOVA detected significant differences in bacterial community composition between fecal samples with low and high composite EED scores using unweighted (R^2^ =0.04, F=2.53, p=0.011) and weighted UniFrac (R^2^ =0.05, F=3.35, p=0.017) (**Figures 6A** and **B**).

**Table 4:** Association between bacterial community composition and individual biomarkers of EED.

| Variable | Unweighted UniFrac |  | Weighted UniFrac |  |
| --- | --- | --- | --- | --- |
| | $R^2$ | p value | $R^2$ | p value |
| **MPO** | 0.02 | 0.07 | 0.01 | 0.6 |
| **AAT** | 0.02 | 0.18 | 0.03 | 0.07 |
| **NEO** | 0.02 | 0.21 | 0.02 | 0.39 |

**Figure 6.**
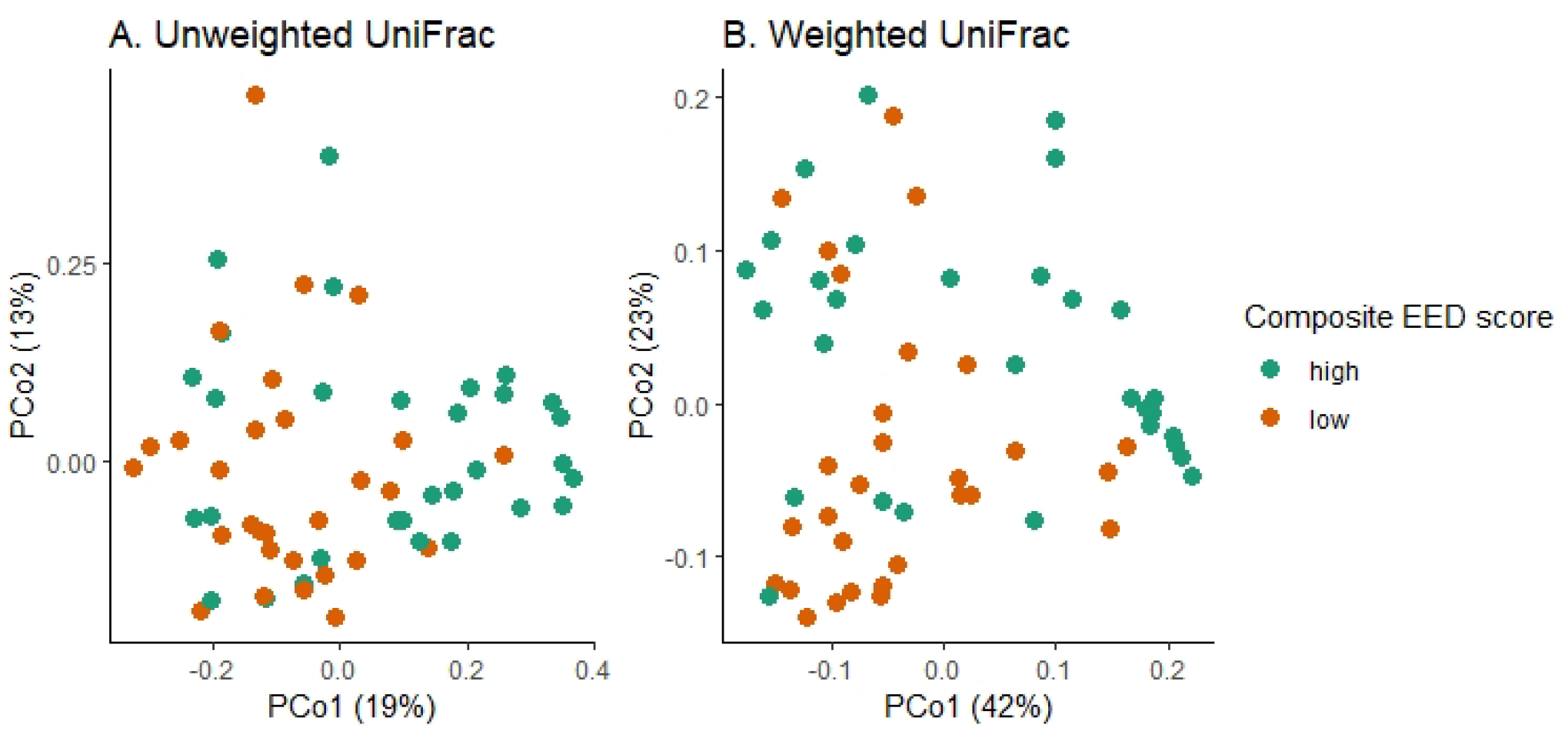
Overall microbiota composition differences by composite EED score. (A) PCoA plot of unweighted UniFrac (R^2^ =0.04, F=2.53, p=0.011) (B) PCoA plot of weighted UniFrac (R^2^ =0.05, F=3.35, p=0.017).

Differential abundance analysis at the genus level, both unadjusted and adjusted for multiple comparison using Benjamin-Hochberg p value correction, was performed to identify genera differentially abundant between low and high EED scores. Three genera, *Succinivibrio, Butyrivibrio* and *Enterococcus*, were differentially abundant in fecal samples with high composite EED score compared to fecal samples with low composite EED score (**Table 5**). An age and sex-adjusted zero-inflated negative binomial regression model was used to further explore this relationship. In the adjusted analysis, only *Succinivibrio* was associated with composite EED score. An increase in the abundance of *Succinivibrio* was associated with 0.14 reduced odds of a having a high composite EED score (**Table 5**).

**Table 5:** Differentially abundant genera in fecal samples with high versus low composite EED scores.

| Genus | log<sub>2</sub>Fold Change | lfcSE | p value<sup>a</sup> | padj<sup>b</sup> | OR<sup>c</sup> (95%CI) | p value<sup>c</sup> |
| --- | --- | --- | --- | --- | --- | --- |
| *Succinivibrio* | -26.3 | 2.6 | <0.001 | <0.001 | 0.14<br>(0.08,0.23) | <0.001 |
| *Butyrivibrio* | -25.6 | 2.9 | <0.001 | <0.001 | 3.52<br>(0.68,18.16) | 0.791 |
| *Enterococcus* | 23.5 | 2.9 | <0.001 | <0.001 | 1.82<br>(0.25,1.2) | 0.619 |

<sup>a</sup>Denotes unadjusted $p$ value while <sup>b</sup>denotes $p$ value obtained after adjusting for multiplicity with Benjamin-Hochberg $p$ value correction.
OR=odds ratio.
CI=confidence interval.
<sup>c</sup>Denotes OR (95%CI) and $p$ values obtained with zero-inflated negative binomial regression (conditional model) after adjusting for age and sex.
lfcSE = log fold change Standard Error.

### Associations between predicted functional metagenome of the microbiota and the composite EED score

We then used PICRUSt2^15^ analysis to predict metagenomes from the 16S sequence data of all 102 baseline fecal samples. This method uses evolutionary modelling to predict which gene families are present and then combines gene families to estimate the composite metagenome and their metabolic pathways. PICRUSt2 analysis predicted that the metagenome of these 102 fecal samples accounted for 431 metabolic pathways, which were collapsed to 67 superpathways for downstream analysis (**Supplementary table S1)**. To explore associations between metabolic pathways identified by PICRUSt2 and the composite EED score, we used the subset of 60 samples that had ELISA results for all the three biomarkers. This sub-set of 60 samples was predicated to contain 52 of the 67 superpathways identified in the 102 sequenced samples, the remaining 15 superpathways were excluded from further analysis. We then used a lasso regression model to determine which of these metabolic pathways were most important for predicting the composite EED score, in which the model coefficient for uninformative variables is reduced to zero. Of the 52 pathways included in the final analysis, the lasso regression model retained five pathways with non-zero coefficients (**Table 6**). We then explored the relationship between these five pathways, and the composite EED score using age-adjusted logistic regression analysis. Three of the five pathways were significantly associated with reduced risk of high EDD scores. A unit increase in the abundance of both the fatty acid biosynthesis and pyrimidine nucleotide degradation pathways was associated with 0.47 reduced odds of high composite EED score while a unit increase in the abundance of tetrapyrrole biosynthesis pathway was associated 0.52 reduced odds of high composite EED score. There were no significant associations between coenzyme B or catechol degradation pathways and the composite EED score in this adjusted analysis (**Table 6**).

**Table 6:** Associations between metabolic pathways and composite EED score.

| Metabolic pathway | <sup>a</sup>Variable importance score | OR (95% CI) | <sup>b</sup>p value |
| --- | --- | --- | --- |
| Pyrimidine nucleotide degradation | 7.80E-05 | 0.47 (0.23,0.81) | 0.006 |
| Fatty Acid biosynthesis | 4.79E-07 | 0.47 (0.23,0.86) | 0.015 |
| Tetrapyrrole biosynthesis | 2.36E-05 | 0.52 (0.27,0.87) | 0.015 |
| Coenzyme B | 7.52E-05 | 0.99 (0.87,1.05) | 0.556 |
| Catechol degradation | 2.43E-04 | 0.99 (0.81,1.25) | 0.982 |

*CI= confidence interval, OR=odds ratio. <sup>a</sup>Pathways with non-zero coefficients retained by lasso regression. <sup>b</sup>Permuted (10,000) p value from an age-adjusted logistic regression.*

## Discussion

We estimated the magnitude of intestinal inflammation and permeability in rural Malawian children using fecal biomarkers and explored associations with the fecal microbiota, characterized by 16S rRNA sequencing. We found elevated fecal levels of NEO in almost two-thirds of the children; MPO was raised in one-third of children whilst AAT was elevated in 10% of children. All biomarker levels decreased with age, consistent with trends shown by previous studies from similar low-income settings. In studies by Naylor *et al*.^6^, Campbell *et al*.^16^ and McCormick *et al*.^12^, more than 80% of infant fecal samples from low income settings had elevated concentrations of MPO, AAT and NEO relative to values from high income settings^6,16^ and all biomarker concentrations decreased with age.

We also found an effect of age on the microbiota. Microbiota diversity was higher in older age groups compared to younger children. Younger children were shown to have a higher abundance of *Actinobacteria* and *Bifidobacterium*, and a lower abundance of the *Bacteroidetes* and *Faecalibacterium* compared to the older age groups. These findings are consistent with trends reported by recent longitudinal studies^17,18^. The increase in diversity and changes in intestinal microbiota seen with the different age groups are also consistent with changes that have been reported during the transition from breast-feeding to weaning^19,20^.

We did not find a relationship between alpha diversity and fecal levels of AAT or MPO, however a marginal negative association was seen between alpha diversity and NEO, a biomarker of intestinal inflammation. To date, there is no other published data available supporting a relationship between fecal microbiota diversity and intestinal inflammation in EED, however, a reduction in fecal microbiota diversity has been reported previously in inflammatory bowel disease^21-23^, which is consistent with our findings. These results suggest a potential role of the gut microbiota in modulating inflammatory responses.

There is limited published data indicating associations between intestinal bacterial community structure and EED. Recently, Ordiz *et al*. found differences in intestinal bacterial composition between children with increased versus normal intestinal permeability as measured by the L:M ratio test^24^; the genera *Succinivibrio, Klebsiella*, and *Clostridium_XI* were less prevalent in children with increased intestinal permeability compared to children with normal levels. Similarly, we found an association between higher abundance of *Succinivibrio* and low composite EED score. *Succinivibrio* is a gram-negative anaerobic commensal of the human colon that has been associated with fiber-degradation^25,26^. The association between *Succinivibrio* abundance and the composite EED score, which is consistent with the results of others^24^, indicates a potential role for *Succinivibrio* in gut function.

Using the 16SrRNA sequencing data generated, PICRUSt2 analysis predicted 67 metabolic pathways of which 5 were identified by the lasso regression model as important for predicting EED. Among the 5 pathways, the fatty acid biosynthesis, pyrimidine nucleotide degradation and tetrapyrrole biosynthesis pathways were significantly associated with composite EED score in an age-adjusted logistic regression model, which to an extent supports the prediction accuracy of the lasso regression model. Fatty acid synthase, an enzyme that catalyzes fatty acid biosynthesis, has been shown to help maintain the intestinal mucus barrier by regulating mucin 2 in murine models^27^. In the present study, we found that an increase in the relative abundance of the fatty acid biosynthesis pathway was associated with reduced chance of high composite EED scores. As such, we can postulate that the fatty acid biosynthesis pathway plays a role in the maintenance of gut integrity in humans. For their part, pyrimidines serve important roles in human metabolism, such as ribonucleotide bases in RNA (uracil and cytosine) and deoxyribonucleotide bases in DNA (cytosine and thymine). Some bacterial species are known to catabolize pyrimidines via a reductive pathway leading to the formation of β-amino acids^28,29^. Dietary supplementation of α-amino acids has been shown to improve intestinal mucosa development in animals^30^, however, there is limited data on the role of β-amino acids formed from the pyrimidine nucleotide degradation pathway in gut function. Thus, the role of the pyrimidine nucleotide degradation pathway in gut function should be explored further. The tetrapyrrole biosynthesis pathway includes pathways in porphyrin (heme) and corrinoid (cobalamin) metabolism. Some porphyrins have been shown to have anti-inflammatory properties through the inhibition of the activity of Fyn, a non-receptor Src-family tyrosine kinase, triggering anti-inflammatory events associated with down-regulation of T-cell receptor signal transduction, leading to inhibition of tumor necrosis factor alpha (TNF-α) production^31^. In the present study, enrichment of the tetrapyrrole biosynthesis pathway was associated with a reduced chance of high composite EED score, a result consistent with the anti-inflammatory properties of components of the tetrapyrrole pathway.

The present study also found a higher abundance of *Succinivibrio* in children with low composite EED scores. This bacterium has been associated with metabolism of nondigestible dietary carbohydrates in humans and animals^25,26^. Among the 67 metabolic pathways predicted by PICRUSt2 in the present study, there were pathways that are involved in carbohydrate metabolism such as the sugar degradation pathway, however these pathways were not identified as being important in predicting EED using the lasso regression model. Further studies are needed to explore the correlations between *Succinivibrio* abundance, carbohydrate metabolism and gut function.

In conclusion, our data suggest that there is an association between intestinal microbiota diversity and intestinal inflammation as measured by fecal NEO. The genus *Succinivibrio*, which was reported by Ordiz *et al*.^24^ to be reduced in EED, was also associated with low composite EED scores in our study. This bacterium has been associated with fiber-degradation^25^ and has recently been shown to be more abundant in children consuming a diet rich in fiber in low-income settings^26^. Future studies should further investigate the role of this bacterium and the predicted metabolic pathways important in EED.

## Limitations of the study

One of the limitations of this study is the use of EED biomarker values from high income countries to define cut-off limits for normal or elevated biomarker concentration. This limitation is not unique to this study; the lack of reference values for the biomarkers of EED in children living in low income settings has been cited by others^6,12^. There is therefore a need for large-scale studies to define reference values of the biomarkers of EED in children living in low income settings. A further limitation was that this was an opportunistic study that used previously collected samples and therefore may not have had adequate statistical power to identify significant associations. A population-based survey would likely have been more robust.

## Resource availability

### Lead contact

Further information and requests for resources should be directed to and will be fulfilled by the lead contact, Martin Holland.

### Materials availability

The study did not generate new unique reagents or materials.

### Data and code availability

The datasets generated from the present study are available from the corresponding author on reasonable request. The V4 16S rRNA sequence data for this study have been deposited in the European Nucleotide Archive (ENA) at EMBL-EBI under accession number PRJEB41238 (https://www.ebi.ac.uk/ena/browser/view/PRJEB41238).

## Methods

All methods can be found in the accompanying Transparent Methods supplemental file.

## Data Availability

The datasets generated from the present study are available from the corresponding author on reasonable request. The V4 16S rRNA sequence data for this study have been deposited in the European Nucleotide Archive (ENA) at EMBL-EBI under accession number PRJEB41238

https://www.ebi.ac.uk/ena/browser/view/PRJEB41238

## Acknowledgments

We would like to acknowledge the support of the field staff at the Blantyre Institute of Community Outreach, Blantyre, Malawi. Funding for this work was provided by the Bill and Melinda Gates Foundation, under grant numbers OPP1066930 and OP1032340.

## Author contributions

Conceptualization, SEB, KK, RLB and MJH; Methodology, DC, HP, and JDH; Formal Analysis, DC and HP; Resources, SEB, KK, RLB and MJH; Data Curation, DC, JDH and KK; Writing – Original Draft Preparation, DC; Writing – Review & Editing, DC, HP, SEB, KMM, KK, RLB and MJH; Supervision, SEB, KK, RLB and MJH; Project Administration; KK and RLB; Funding Acquisition, SEB, KK, RLB and MJH. All authors read and approved the final manuscript.

## Supplemental Information

## Transparent methods

### Study design

This study utilized baseline samples from a nested cohort study, conducted within the framework of a randomized controlled trial (NCT02047981). The study design and methodology for sample selection of both the nested cohort and the parent trial have been previously described^14,32,33^. Within the nested cohort study, 102 baseline samples from children residing in 30 clusters within the Mangochi district were analysed by 16S rRNA gene sequencing. All sampled children were aged between 1 and 59 months and weighed at least 3.8kg. Sample collection took place between May and July 2015.

### Fecal sample collection

Fecal samples for participating children were collected by their mothers or guardians, who were provided with sterile fecal collection bottles (Wheaton, UK) and given verbal instruction, by a nurse, on how to collect the sample. Fecal samples were returned to the field team as soon as possible after collection. These samples were put in a cool box with ice packs until the end of the day (not more than 8 hours after collection) and were brought to the laboratory where they were placed at -80°C for storage.

### Fecal biomarkers of EED

Commercially available ELISA kits for MPO (Immundiagnostik, Germany), AAT (Immundiagnostik) and NEO (Genway, CA, USA) were used for the quantification of each biomarker. Each assay was performed according to the kit instructions using stool diluted 1/500 for MPO, 1/25,000 for AAT and 1/100 in phosphate buffered saline (PBS) for NEO^12^. Fecal samples with out-of-range concentration values were re-tested at appropriate dilutions until in-range values were obtained.

### 16S rRNA gene sequencing and sequence processing

16S rRNA gene sequencing and sequence processing were performed as detailed elsewhere^14^. Briefly, DNA libraries were prepared by amplifying an approximately 390-bp V4 region of the 16S rRNA gene in the genomic DNA extracted from 250mg of fecal sample. The amplicons were purified and then quantified on a Qubit 2.0 Fluorometer (Invitrogen) with high-sensitivity reagents (Life Technologies) and thereafter pooled in equimolar amounts. A PhiX-spiked DNA library was then assayed by 2×300bp paired-end sequencing on the Illumina MiSeq platform for a total of 600 cycles using a standard protocol^34^.

Raw sequence data was processed in QIIME 2^35^ as per standard practice^34^ and *de novo*-clustered into operational taxonomic units (OTUs) at ≥ 97% identity. Taxonomy was assigned to the OTUs using a naïve Bayes classifier pre-trained on the SILVA 16S database^36^ subsequently generating an OTU table. To exclude spurious OTUs, only bacterial OTUs identified to the genus level, with sequences more than 0.005% of the total number of sequences^37^ and a frequency of more than 0.01% in any sample were retained in the analyses.

### Statistical analysis

Shannon (H) and Simpson (D) diversity indices were calculated from the OTUs to indicate alpha diversity using the phyloseq R package^38^. The differences in Shannon and Simpson distribution between groups were statistically tested using ANOVA and the non-parametric Kruskal-Wallis test respectively. Weighted and unweighted UniFrac distance matrices were also calculated to determine bacterial phylogenetic distance between samples.

Median (25^th^, 75^th^ quartile) concentration of individual fecal biomarkers of EED were used to determine their distribution. Published cut-off values for each of the 3 biomarkers^12,13^ were used to determine proportions of fecal samples with elevated biomarkers. Concentrations of the 3 biomarkers were combined to form a composite EED score following the method of Kosek *et al*.^13^ with the following modification: absolute weightings instead of rounded weightings from the principal component analysis^13^ were used to generate continuous EED scores.

Associations between alpha diversity indices and biomarker concentrations were investigated using regression analyses adjusted for age and sex. PERMANOVA with 1000 permutation tests was run on weighted and unweighted UniFrac distances to compare bacteria community compositional differences between groups; differences in UniFrac clustering by groups were visualized using principal coordinate analysis (PCoA) plots. Differential abundance analysis at the genus level was performed to identify OTUs differentially abundant in EED using the DESeq2 package in R^39^. Statistical significance of log_2_ fold changes was assessed using the default Wald test with Benjamin-Hochberg p value correction. Cut-off for all significant tests was set at p<0.01. Zero-inflated negative binomial regression analysis was conducted, adjusting for age and sex, to establish the relationship between the differentially abundant OTUs and the composite EED score.

PICRUSt2^15^ analysis was used to predict metagenomes from 16S data and a reference genome database. We included OTU abundance data from 102 baseline samples that passed previously described quality control steps. Briefly, unique sequences representing the previously defined OTUs were added to reference multiple-sequence alignments and phylogeny containing 20000 full 16S rRNA genes of archaea and bacteria from the IMG database. Hidden-state prediction based on position of OTU sequences in the reference phylogeny was used to predict gene family abundances. To estimate functional profiles of each sample, gene family abundances per OTU were adjusted for OUT abundance values per sample. Sample functional profiles were finally utilized to infer MetaCyc pathway abundances. PICRUSt2 identified 431 metabolic pathways; these were collapsed to 67 superpathways for downstream analysis. To explore associations between metabolic pathways identified by PICRUSt2 and EED, only the 60 samples that had ELISA results for all the three biomarkers from which we were able to calculate the composite EED score were included. In this subset of 60 samples, 15 super pathways had zero count score contribution and were excluded from further analysis. Lasso regression was then used to identify the inferred metabolic or functional properties of 16S rRNA community that are important for EED, the tuning parameter (lambda) was chosen by ten-fold cross-validation. Age-adjusted logistic regression analysis with permuted p values was used to explore associations between pathways predicted by PICRUSt2, and identified as important by the lasso regression model, and the composite EED score.

### Ethical considerations

This study was conducted in accordance with the Declaration of Helsinki. It was approved by the London School of Hygiene and Tropical Medicine Ethics Committee (UK) and the College of Medicine Research Ethics Committee (Malawi). Information and consent forms were translated into local languages (Yao and Chichewa) prior to their approval by the local ethics committee. Consent was first obtained at the community level through discussions with the village chief and community elders who then indicated the willingness, or unwillingness, of the community to participate through verbal consent. Written, informed consent (by thumbprint or signature) was then obtained from the parent or legal guardian of each child for inclusion before they participated in the study. During the consenting process, all parents and guardians were informed of their freedom to withdraw their child from the study at any time without giving reason for doing so.

**Supplementary table S1:**
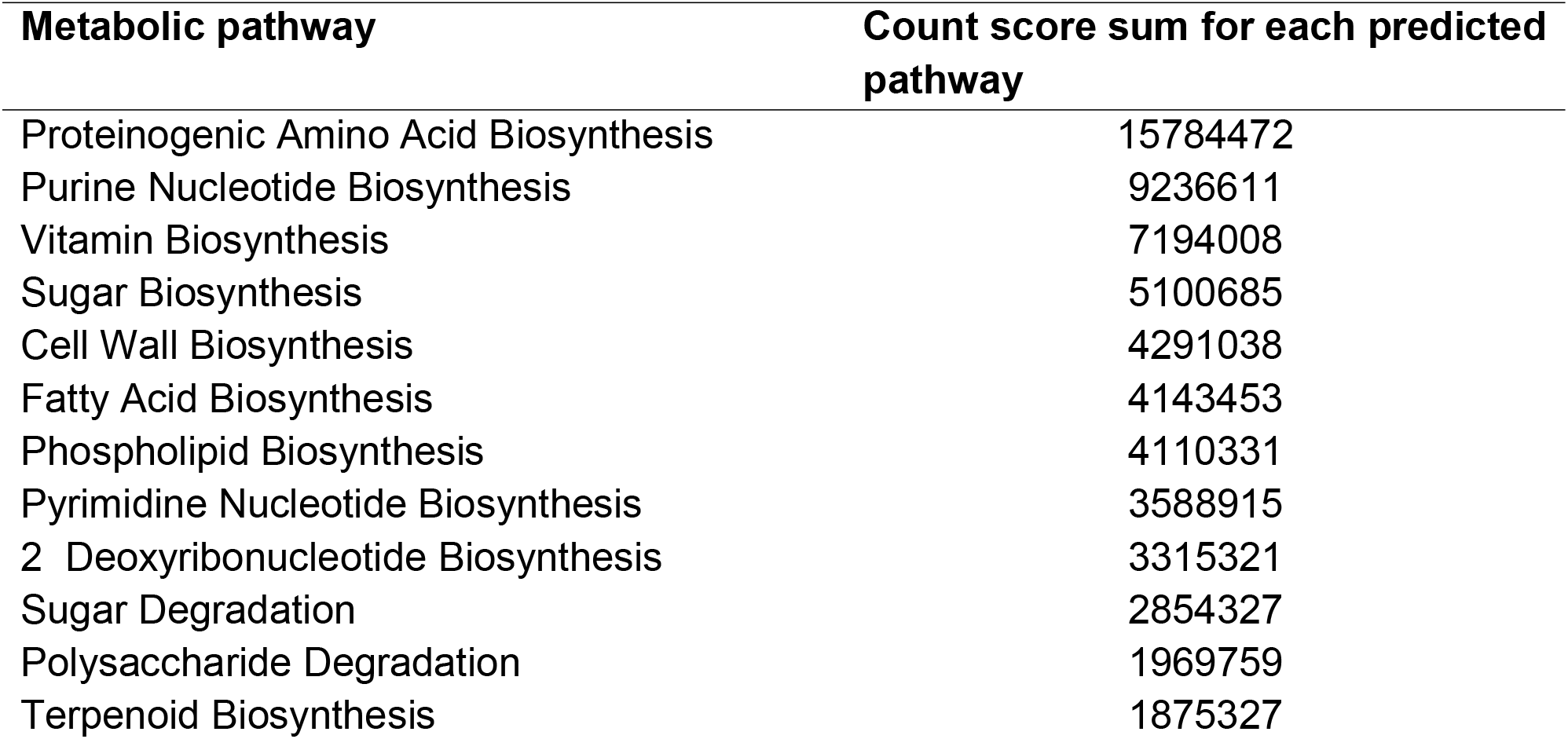
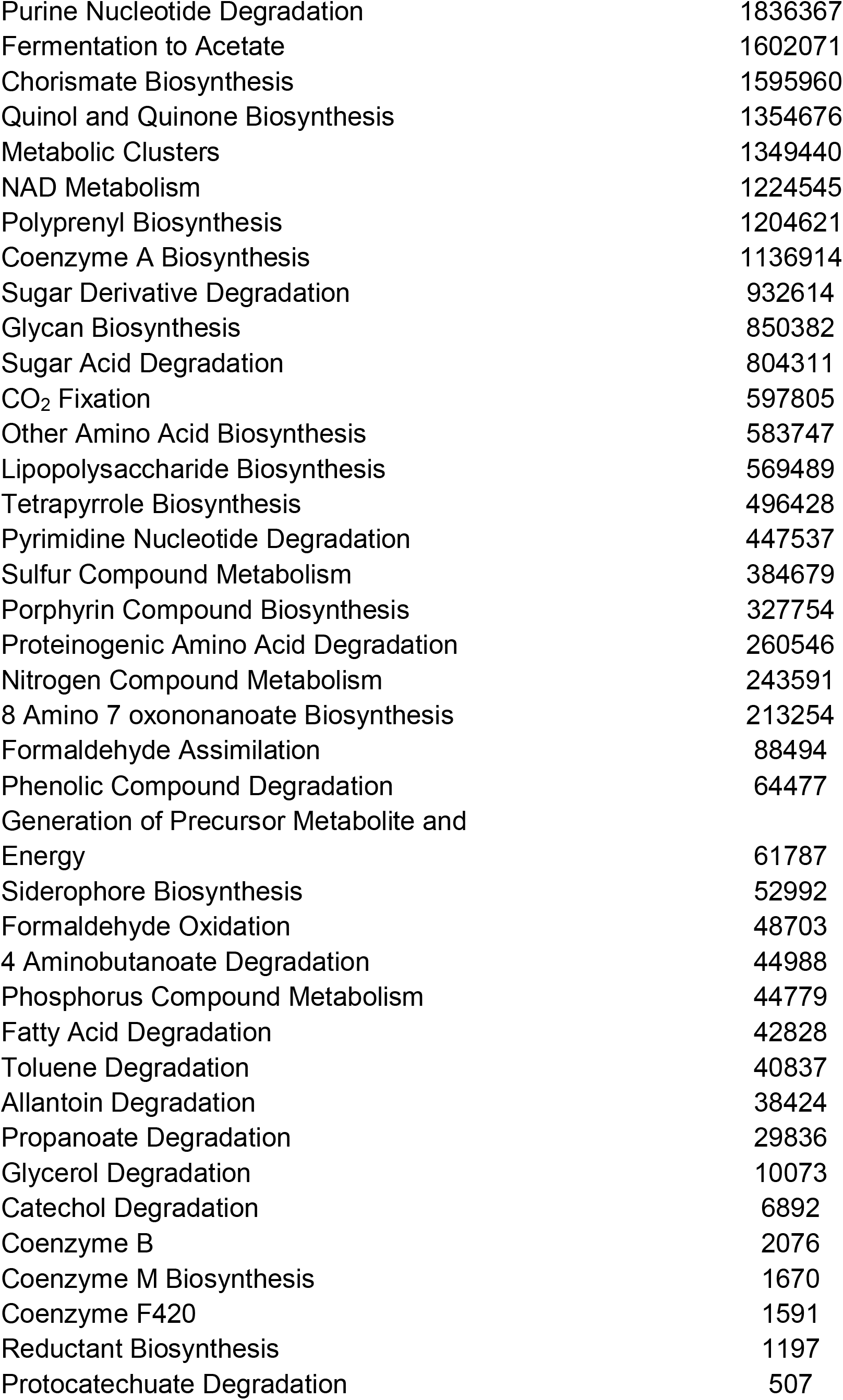
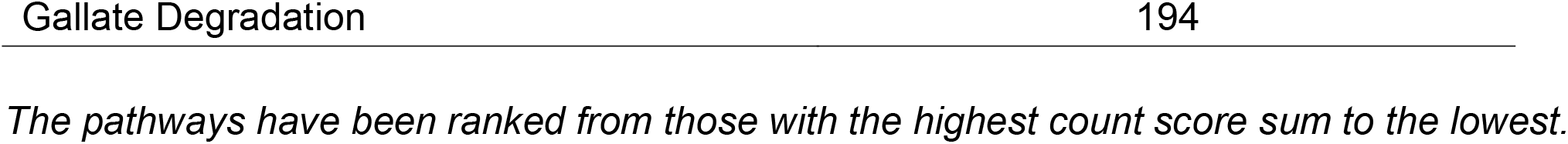
Metabolic pathways predicted by PICRUSt2.

